# Targeted MRM-analysis of plasma proteins in frozen whole blood samples from patients with COVID-19

**DOI:** 10.1101/2023.09.20.23295832

**Authors:** Anna E. Bugrova, Polina A. Strelnikova, Alexey S. Kononikhin, Natalia V. Zakharova, Elizaveta O. Diyachkova, Alexander G. Brzhozovskiy, Maria I. Indeykina, Ilya N. Kurochkin, Alexander V. Averyanov, Evgeny N. Nikolaev

## Abstract

The COVID-19 pandemic has exposed a number of key challenges that need to be urgently addressed. In particular, rapid identification and validation of prognostic markers is required. Mass spectrometric studies of blood plasma proteomics provide a deep understanding of the relationship between the severe course of infection and activation of specific pathophysiological pathways. Analysis of plasma proteins in whole blood may also be relevant for the pandemic as it requires minimal sample preparation. Here, for the first time, frozen whole blood samples were used to analyze 189 plasma proteins using multiple reaction monitoring (MRM) mass spectrometry and stable isotope-labeled peptide standards (SIS). A total of 128 samples (FRCC, Russia) from patients with mild (n=40), moderate (n=36) and severe (n=19) COVID-19 infection and healthy controls (n=33) were analyzed. Levels of 114 proteins were quantified and compared. Significant differences between all of the groups were revealed for 61 proteins. Changes in the levels of 30 reproducible COVID-19 markers (SERPING1, CRP, C9, ORM1, APOA1, SAA1/SAA2, LBP, AFM, IGFALS, etc.) were consistent with studies performed with serum/plasma samples. Levels of 70 proteins correlated between whole blood and plasma samples. The best-performing classifier built with 13 significantly different proteins achieved the best combination of ROC-AUC (0.93-0.95) and accuracy (0.87-0.93) metrics and distinguished patients from controls, as well as patients by severity and risk of mortality. Overall, the results support the use of frozen whole blood for MRM analysis of plasma proteins and assessment of the status of patients with COVID-19.

## 1. Introduction

The COVID-19 pandemic has revealed some important aspects that require urgent decision-making and has compelled the global community to rapidly develop both effective therapeutic approaches and diagnostic methods, including those that predict the risk of an adverse outcome. The lessons of this pandemic must certainly be learned in order to effectively respond to other similar challenges, and they concern not only urgent innovations, but also the significantly increased workload for medical institutions and analytical laboratories due to the high influx of patients and samples for analysis. Particularly, mass spectrometry (MS)-based proteomics has the potential to be used as an ideal analytical technology in such situations, as it can provide the fastest, deepest unbiased analysis necessary for the understanding of the role of specific biological processes in the ongoing pathophysiological changes, as well as to create specific marker panels [1,2]. For productive analysis of the increased number of samples, it is also essential to use the least time-consuming sample preparation methods to minimize erroneous results.

Blood serum and plasma are among the most traditional samples for clinical assays and have been used in the largest number of biomarker studies of COVID-19. In the case of this infection, blood studies are of particular relevance because the coronavirus affects the functioning of the capillary endothelium by promoting its inflammation and can cause acute distress respiratory syndrome, multiple organ dysfunction or even sepsis [3-6], and thus undoubtedly affects the lipidomic, proteomic and metabolomic plasma/serum profiles [7-10]. As early as in July 2020, several untargeted high-resolution MS based proteomic studies of blood serum were performed and showed good intersections with each other in a number of dysregulated proteins and, in general, complemented each other in the list of potential markers of disease severity [11-13]. Already these first results indicated the relationship between the severe course of disease progression and elevated levels of coagulation and complement components [13], as well as with activation of acute phase proteins and down-regulation of some apolipoproteins accompanied by dysregulation of metabolites involved in lipid metabolism [11]. One way or another, all further untargeted and targeted MS studies of serum and blood plasma, with or without depletion of major proteins, led to similar conclusions [14-24]. Overall, ∼100 proteins have been shown in at least two different MS studies to be concordantly dysregulated. For 15 of them, concordant dysregulation has been shown in at least five studies. These include 10 up-regulated proteins: alpha-1-antichymotrypsin (SERPINA3) [11,14-23], plasma protease C1 inhibitor (SERPING1) [11,13,17-19,21-23], C-reactive protein (CRP) [11,12,14,15,21-23], complement component C9 (C9) [11,13,16,18,22,23], alpha-1-acid glycoprotein 1(ORM1, AGP1) [14,16-18,22,23], von Willebrand factor (VWF) [16,17,19,21-23], inter-alpha-trypsin inhibitor heavy chain 3 (ITIH3) [11,12,16,18,22], actin (ACTB, ACTC1, ACTA2) [12,14,21-23], serum amyloid A-1 and A2 proteins (SAA1, SAA2) [11,12,14,22,23], lipopolysaccharide-binding protein (LBP) [11,12,17,22,23]; as well as 5 down-regulated proteins: histidine-rich protein (HRG) [15,18-22], apolipoprotein A-I (APOA1) [12,16,18,21,22], afamin (AFM) [14,17,18,21,22], insulin-like growth factor-binding protein complex acid labile subunit (IGFALS) [14,20-22,24], and N-acetylmuramoyl-L-alanine amidase (PGLYRP2) [11,14,18,21,22].

Whole blood is not such a popular research object due to its much higher proteome complexity and domination of red blood cell proteins. However, the possibility of serum/plasma preparation may be more or less limited under the extreme circumstances of a pandemic with a significantly increased influx of hospitalized patients. At the same time, whole blood samples do not require even the simplest processing, small samples can be collected even by the patient at home, and can easily be stored frozen or as dried blood spots. Therefore, for pandemic conditions, it seems particularly appropriate to assess the diagnostic potential of whole-blood proteomics.

Until now, studies of plasma proteins in whole blood have not been very popular. This may be due to the predominant use of immuno-based approaches for assays targeted at specific proteins. While, the use of complex subjects such as whole blood greatly increases the number of non-specific and false positive results. There are two ways to process and store whole blood: drying or freezing the sample. Usually for MS-based proteomic analysis the dry blood spots (DBS) technique (when blood samples are blotted and dried on filter paper) is used [25-27]. The use of volumetric absorptive microsampling (VAMS) to deplete highly abundant proteins allows to reach ∼2000 protein identifications in DBS [28]. But frozen whole blood samples are much easier and faster to collect and are commonly used for genetic or immunological studies [29]. Therefore, their additional proteomic analysis may be particularly relevant in a pandemic. However, quantitative MS studies of plasma proteins in frozen whole blood are rare and no such studies have been performed during the COVID-19 pandemic.

In this study, the ability of MRM-MS to quantify plasma proteins in frozen whole blood samples was evaluated. Samples collected during the COVID-19 pandemic from 95 patients with varying disease severity and a group of healthy controls were analyzed to estimate the levels of 189 blood plasma proteins using stable isotope-labeled peptide standards (SIS). The analyzed proteins included ∼100 previously reported COVID-19 markers proposed in previous MS studies with blood plasma and sera. A combination of statistics and machine learning was used for data analysis in order to build the best-performing classifier for severity prediction in patients with COVID-19.

## 2. Materials and Methods

### 2.1 Study population

Participants were recruited in the Department for treatment of the novel coronavirus infection at the Federal Research Clinical Center (FRCC) under Federal Medical and Biological Agency (Russia) from 25.06.2021 till 19.07.2021. 95 patients were positive according to PCR-testing for SARSCoV-2. All 95 patients have been hospitalized; all of them had viral pneumonia confirmed by high resolution computer tomography of the chest. The group of healthy controls (n=33) was recruited among the FRCC staff. Informed consent was obtained from all participants, and the study was approved by the FRCC local ethical committee (clinical protocol No. 5, 11 May 2021). Patients were divided into severity groups according to the respiratory support they needed. Mild (n=40) -did not need oxygen supply, moderate (n=36) needed low flow oxygen support, and severe (n=19) needed high flow nasal oxygen, non invasive or invasive mechanical ventilation (11 of them died). The demographic and clinical characteristics of the studied groups are presented in Table 1.

**Table 1.** Subject demographics.

|  | COVID-19 patients |  |  |  | Controls |
| --- | --- | --- | --- | --- | --- |
|  | Total | Mild | Moderate | Moderate |  |
| N | 95 | 40 | 36 | 19 | 33 |
| Age (years)<sup>a</sup> | 55 (44; 70) | 50 (37; 63) | 61 (52; 69) | 67 (52; 73) | 57 (41; 68) |
| Sex (% , F) | 57 | 63 | 55 | 44 | 67 |
| Body mass index<sup>a</sup> | 27 (25; 33) | 25 (24; 29) | 29 (26; 34) | 29 (25; 39) | 26 (22; 31) |
| Smokers (%) | 17 | 9 | 18 | 34 | 30 |
| Onset of symptoms (days)<sup>a</sup> | 6 (4; 8) | 7 (5; 8) | 6 (4; 8) | 6 (4; 7) | - |
| Hospitalization duration (days)<sup>a</sup> | 10 (7; 14) | 7 (6; 9) | 11 (9; 14) | 15 (13; 18) | - |
| Lung damage (% , CT) | 30 (25; 50) | 25 (15; 25) | 40 (25; 50) | 75 (62; 75) | - |
| CRP (mg/L)<sup>a</sup> | 43 (14; 70) | 22 (10; 50) | 58 (17; 99) | 47 (19; 99) | 3.5 (1.8; 12.4) |
| Total protein (g/L)<sup>a</sup> | 69 (64; 73) | 68 (65; 73) | 70 (67; 73) | 63 (61; 70) | 73 (68; 74) |
| Leukocytes (10<sup>9</sup>/L)<sup>a</sup> | 5 (4; 6) | 4 (4; 6) | 5 (4; 7) | 5 (5; 6) | 8.6 (6.5; 10.7) |
| ESR (mm/hour)<sup>a</sup> | 33 (20; 46) | 26 (18; 40) | 40 (28; 60) | 34 (24; 46) | 38 (21; 60) |
| Lymphocytes (%) | 21 (14; 28) | 27 (21; 32) | 18 (13; 23) | 16 (11; 20) | 27 (17; 32) |
| Lymphocytes (10<sup>9</sup>/L)<sup>a</sup> | 1.0 (0.7; 1.4) | 1.2 (0.9; 1.4) | 1.0 (0.6; 1.4) | 0.9 (0.6; 1.1) | 2.0 (1.5; 2.6) |

<sup>a</sup> - Median values are given; the 0.25 and 0.75 percentiles are indicated in brackets.
Abbreviations: CT - computed tomography scan; CRP - C-reactive protein; ESR - erythrocyte sedimentation rate.

### 2.2. Whole blood sample collection and preparation for MS

Venous blood was collected using vacuum tubes with K^2+^-EDTA, aliquoted and stored at -80 °C. The study was performed using isotope-labeled standard (SIS) “heavy” peptides, which were added to each sample and acted as internal standards for normalization, and unlabeled “light” peptides (NAT), which were used to create quantitative calibration curves. Synthesis and characterization of SIS and NAT peptides was carried out in the Omics lab at Skoltech using standard procedures, which were previously described in detail [23,24,30]. The list of peptides and proteins is provided in Supplementary Table S1.

Sample preparation was carried out using 10 μL of whole blood, similar to the protocols applied for plasma [23,24,31]. After thawing, the blood samples were carefully mixed and centrifuged at 4000g for 10 minutes. Before trypsinolysis, the samples were denaturated and reduced by incubation with 6 M urea, 13 mM dithiothreitol and 200 mM Tris × HCl (pH 8.0, +37 °C, 30 min). Next, the proteins were alkylated by a 30-min incubation in the dark with 40 mM iodoacetamide. For trypsinolysis, the samples were diluted with 100 mM Tris × HCl (pH 8.0) until <1 M urea; L-(tosylamido-2-phenyl) ethyl chloromethyl ketone (TPCK)-treated trypsin (Worthington) was added at a 20:1 (protein:enzyme, *w*/*w*) ratio; and the samples were incubated for 18 h at 37 °C. The reaction was quenched by acidifying the reaction mixture with formic acid (FA) to a final concentration of 1.0% (pH ≤ 2), and an estimated peptide concentration of ∼1 mg/mL [30]. Then each sample (40 μL) was spiked with 10μL of the SIS peptide mixture, prepared by solubilization in 30% ACN/0.1% FA and dilution to 10× LLOQ per μL with 0.1% FA. All samples were then concentrated by solid-phase extraction (SPE) using an Oasis HLB μElution plate: 1) the plate was conditioned with MeOH (600μL), equilibrated with 0.1% FA (600μL), and loaded with samples; 2) the wells were washed with H_2_O (600μL, ×3); 3) bound peptides were eluted with 70% ACN/0.1% FA (55 μL) [24]. For each reference standard and quality control sample, 40 μl of a BSA surrogate matrix digest (143 μg/mL) was spiked with the SIS peptide mixture, as well as with the same volume of a level-specific “light” peptide mixture at a ratio of 4:1:1 (v/v/v). The standard curve and quality control samples were subjected to the same SPE procedure. All SPE eluates were evaporated using a speed vacuum concentrator and stored at -80°C. Prior to LC-MS/MS analysis, the samples were reconstituted in 34 μL of 0.1% FA.

### 2.3. Plasma samples

Plasma samples from 32 healthy controls were prepared in parallel with their whole blood samples to study the correlation between target protein concentrations. Plasma was obtained by centrifuging the K^2+^-EDTA whole blood samples at 4000 × *g* for 10 min at room temperature within 1h after collection. The aliquots were stored at -80°C. Sample preparation for MRM-MS was performed as described above for whole blood samples.

### 2.4. LC-MS/MS analysis and MS data processing

All samples were analyzed in duplicate by HPLC-MS using an ExionLC™ UHPLC system (Thermo Fisher Scientific, Waltham, MA, USA) coupled online to a SCIEX QTRAP 6500+ triple quadrupole mass spectrometer (SCIEX, Toronto, ON, Canada). LC and MRM parameters were adapted and optimized basing on the previous studies done with the BAK125/270 kits [30, 31].

The loaded sample volume was 10 μL per injection. HPLC separation was carried out using an Acquity UPLC Peptide BEH column (C18, 300 Å, 1.7 μm, 2.1 mm × 150 mm, 1/pkg) (Waters, Milford, MA, USA) with gradient elution. Mobile phase A was 0.1% FA in water; mobile phase B -0.1% FA in acetonitrile. LC separation was performed at a flow rate of 0.4 mL/min using a 53-min gradient from 2 to 45% of mobile phase B. Mass spectrometric measurements were carried out using the MRM acquisition method. The electrospray ionization (ESI) source settings were as follows: ion spray voltage 4000 V, temperature 450°C and ion source gas 40 L/min. The corresponding transition list for MRM experiments with retention time values and Q1/Q3 masses for each peptide is available in Table S1.

For quantitative analysis of the LC-MS/MS raw data, Skyline Quantitative Analysis software (version 20.2.0.343, University of Washington) was used. To calculate the peptide concentrations in the measured samples (fmol per 1 µL of plasma), calibration curves were generated using the 1/(x2)-weighted linear regression method. All experimental results were uploaded to the PeptideAtlas SRM Experiment Library (PASSEL) and are available via link: http://www.peptideatlas.org/PASS/PASS05842 (accessed on 4 September 2023).

### 2.5. Statistical Analysis

Statistical analysis and data visualization were performed using Python (3.7.3) scripts with SciPy [32], Seaborn [33], matplotlib [34], and Pandas [35] packages.

Proteins identified in less than 60% of the samples were excluded; the final number of analyzed features was 114. The data was Log_2_(x+1) transformed. “NaN” -values were replaced with random non-zero values using a Gaussian distribution with a shift down = 0.4 and width = 0.2 of the mean value for each group of patients (control, mild, moderate, and severe).

The Mann-Whitney test was used to evaluate statistical differences between the groups. p-values were adjusted using the Benjamini-Hochberg method. The Cohen’s size (mean difference divided by the variance for different groups of samples) was also considered.

The ordinary least squares (OLS) method was used to construct the linear regression model in order to compare the data from paired whole blood and plasma samples from control patients.

### 2.6. Machine Learning

The Scikit-Learn package was used to obtain all machine learning models [36]. To differentiate between pairwise groups, 4 classifying algorithms were considered: k-Nearest Neighbors (kNN), Logistic Regression (LR), Random Forest (RF), and the Support Vector Classifier (SVC). For the application of all algorithms, except RF, normalized data were used. The best models were determined using k-Fold cross-validation (k = 4). Features were selected according to preliminary statistical analysis. Specific hyper-parameters for each algorithm were selected using a grid search based on the highest ROC-AUC scores for different combinations of compared groups and protein marker panels. The proposed hyper-parameters for the classifiers discussed in the “results” section are presented in the Supplementary materials (Table S5).

## 3. Results

### 3.1. Significantly dysregulated plasma proteins in whole blood

In total, 189 target plasma proteins were analyzed in whole blood samples using LC–MRM MS with corresponding SIS peptides and resulted in 114 quantitatively measured proteins. (Supplementary Table S2). These 114 proteins included 89 previously described COVID-19 markers, with 54 reproducible markers according to ≥2 studies, of which 36 were consistently dysregulated in ≥3 different studies and 10 were shown to be significant in ≥5 studies.

Pairwise comparison of different groups of patients with each other and with healthy controls revealed a very large number of proteins with statistically significant differences (Supplementary Table S3). A total of 61 proteins passed the 5% FDR cutoff after the Benjamini–Hochberg multiple testing correction in at least one pairwise comparison: 21 were up-regulated, 36 were down-regulated, and 4 showed different directions for different comparison groups (Table 2).

**Table 2.**
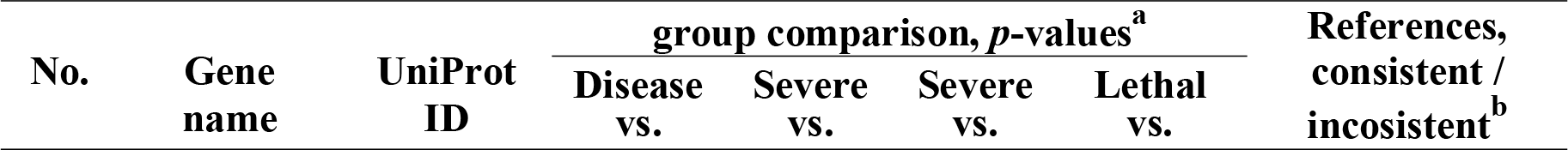
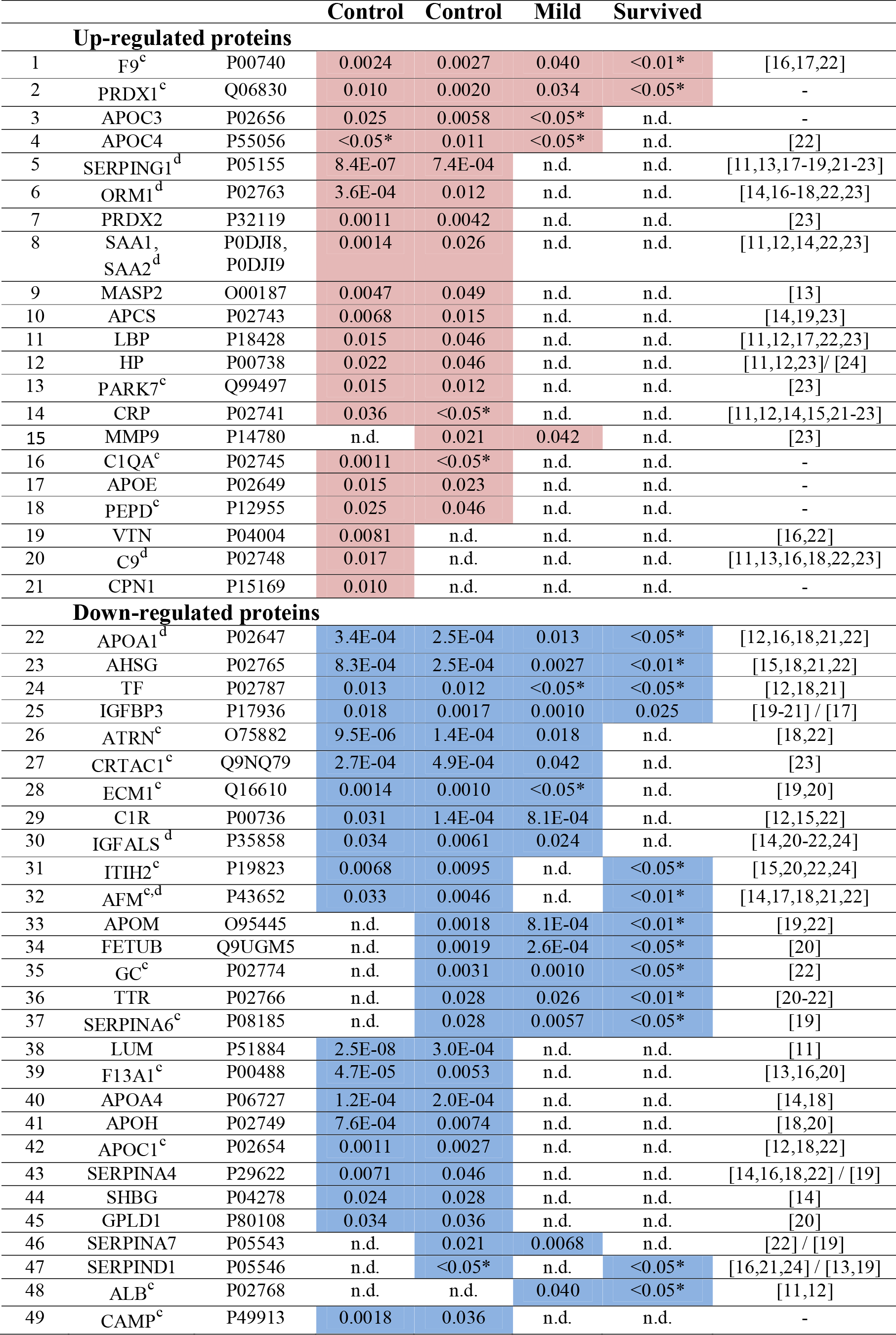
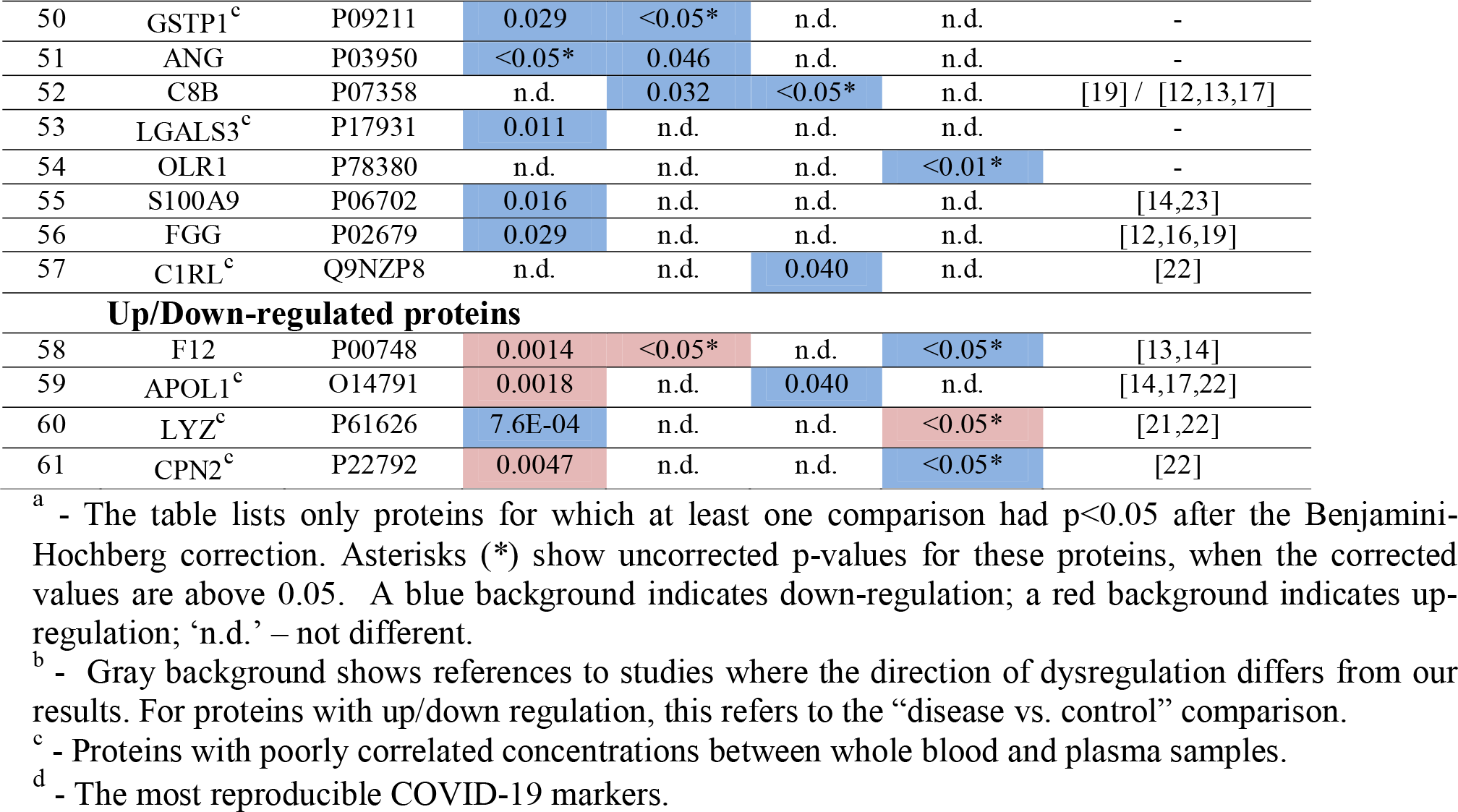
Significantly dysregulated plasma proteins in whole blood.

Six of the revealed significantly different proteins (up-regulated F9 and PRDX1, and down-regulated APOA1, AHSG, TF and IGFBP3) deserve special attention as they were found to be different between all groups presented in Table 2. Thus, they not only distinguish patients from healthy controls but also distinguish patients by severity and risk of mortality. Nevertheless, some other proteins (including SERPING1, ATRN, LUM, and ORM1) demonstrate an even greater difference between groups of patients and healthy controls (Fig. 1).

**Figure 1.**
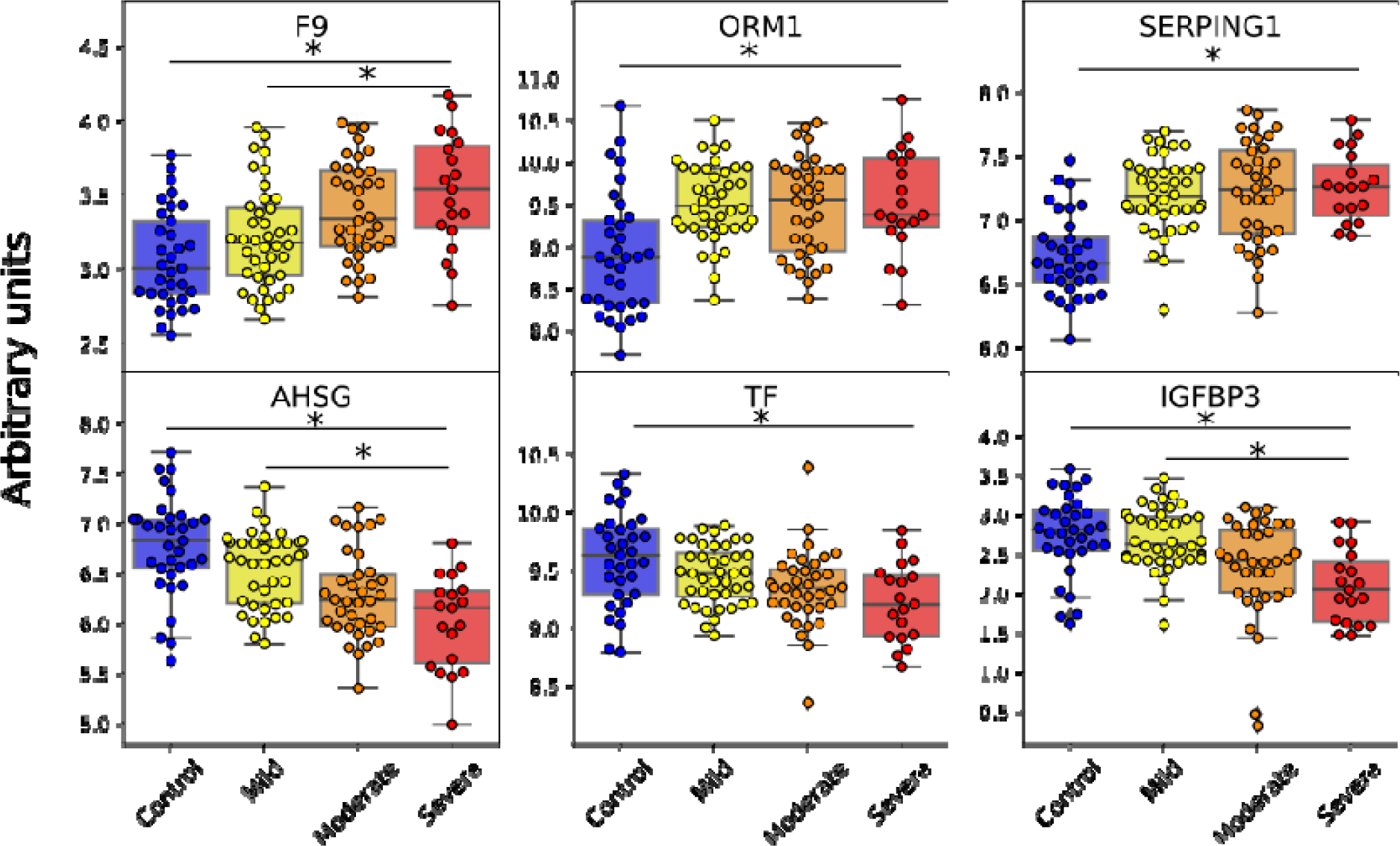
Significantly different proteins that distinguish various groups. Asterisks (*) indicate a significant difference at *p*<0.05 after the Benjamini–Hochberg correction. Protein levels are shown in arbitrary units on a Log_2_(x+1) scale.

In general, the results presented in Table 2 show very good agreement with the results of other studies and a very insignificant number of inconsistencies. It is noteworthy that most literary data concerns serum analyses [12-15] and research that involves the depletion of major plasma proteins [19,22]; nevertheless, for most proteins, our findings still align well with these studies. Notably, 9 of the analyzed top 10 COVID-19 markers also showed significant differences in whole blood samples, while no significant difference was found only for ACTA2 (Supplementary Table S3).

### 3.2. Correlation of protein concentrations between whole blood and plasma

To better understand the relevance of comparing results obtained on whole blood with published results for plasma/serum, analysis of the correlations is particularly important. For this analysis of paired whole blood and plasma samples from 32 healthy controls was carried out. The constructed linear regression for the average protein concentrations confirms a linear relationship between measurements in whole blood and plasma (Fig. 2). Overall, the data are well described by the proposed linear model, with 93% of the proteins falling into the 95% prediction interval with only 8 outliers. Even taking into account the data for all proteins, the coefficient of determination R^2^ turned out to be 0.58 (>0.5), what suggests a good fit of the model to the data. After removing the 8 outliers the R^2^ coefficient increased to 0.88, indicating a much better agreement between the data and the constructed linear model.

**Figure 2.**
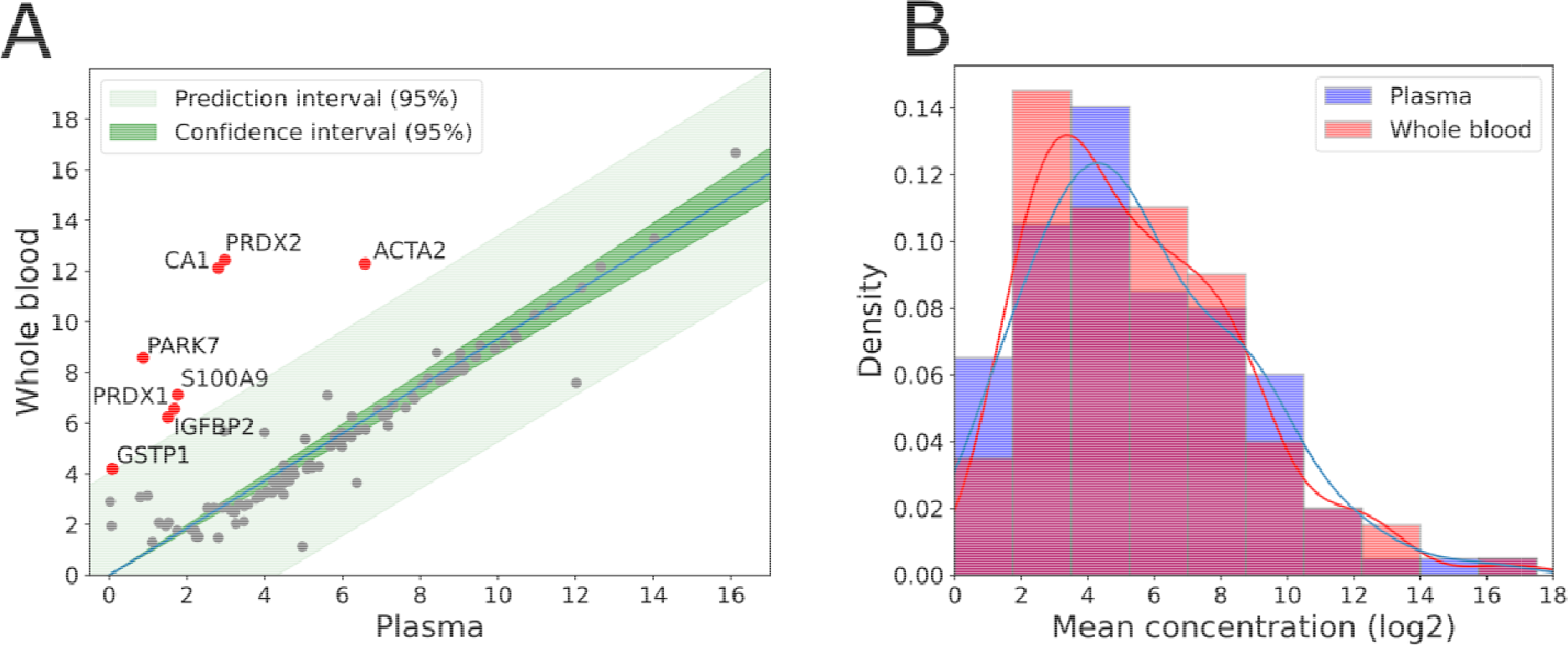
Comparison of target protein concentrations between paired frozen whole blood and plasma samples from 32 healthy controls. (A) Linear regression plot of average protein concentrations. Red dots indicate proteins that fall outside the predicted interval (95%) for the constructed linear regression model. (B) Distribution profiles of average protein concentrations in plasma and whole blood samples.

To assess the coherence of plasma-blood data for each individual protein Pearson correlation coefficients for paired plasma and blood samples were estimated (Supplementary Table S4). According to these calculations, 19 proteins showed a strong positive correlation (R>0.8, *p*-value <0.001), 17 proteins were moderately correlated (R>0.7, *p*-value <0.001), and 34 were weakly correlated (R>0.35, *p*-value <0.05). The non-correlating proteins (R<0.35, *p*-value >0.05) included 22 of the 61 significantly different proteins mentioned in Table 2. Most of the outliers also turned out to be non-correlating: ACTA2, PARK7, PRDX1, IGFBP2, and GSTP1.

## Building classifiers distinguishing between patients and healthy controls

Differences in the proteomic profiles among severity groups can be used to identify unique features for COVID-19 stage classification with a machine-learning approach. Four algorithms were considered to build a classifier: logistic regression (LR), random forest (RF), k-nearest neighbors (kNN) and the support vector classifier (SVC) (Supplementary Table S6).

Initially 19 proteins significantly different between the mild and severe COVID-19 groups were considered as a “reference” set: C1R, APOM, FETUB, IGFBP3, GC, AHSG, SERPINA6, SERPINA7, APOA1, ATRN, IGFALS, TTR, APOL1, PRDX1, C1RL, ALB, F9, CRTAC1, and MMP9 (Table 2).

Features that were significant for several pairwise comparisons seemed to be of particular importance (F9, PRDX1, APOA1, AHSG, TF, and IGFBP3). While, proteins that did not show differences between controls and patients were omitted from the list (APOM, FETAB, GC, SERPINA6, SERPINA7, TTR, C1RL, ALB, and MMP9). In addition, APOL1 was also excluded due to its opposite regulation in different pairwise comparisons (Table 2). Therefore, the first proposed panel consisted of just 9 proteins including с1R, IGFBP3, AHSG, APOA1, ATRN, IGFALS, PRDX1, F9, and CRTAC1. An LR-based classifier reached the highest ROC-AUC (0.97) and accuracy (0.90) characteristics for this panel for distinguishing between mild and severe COVID-19 groups. For determination of lethal cases, the best metrics were shown with the kNN classifier (ROC-AUC = 0.94, accuracy = 0.93). However, none of the classifiers obtained with this panel reached the 0.9 threshold of the ROC-AUC metric in other pairwise comparisons (Table 3).

**Table 3.** ROC-AUC and accuracy characteristics of different classifiers.

| Panel | Algorithm | ROC-AUC / Accuracy |  |  |
| --- | --- | --- | --- | --- |
|  |  | Severe vs. Mild | Disease vs. Control | Lethal vs. Survived |
| **9 proteins:**<br>C1R, IGFBP3, AHSG, APOA1, ATRN, IGFALS, PRDX1, F9, CRTAC1 | LR | 0.98 / 0.90 | 0.89 / 0.84 | 0.90 / 0.79 |
|  | RF | 0.96 / 0.86 | 0.85 / 0.80 | 0.92 / 0.93 |
|  | kNN | 0.96 / 0.88 | 0.82 / 0.81 | 0.94 / 0.93 |
|  | SVC | 0.95 / 0.86 | 0.87 / 0.78 | 0.89 / 0.93 |
| **12 proteins:**<br>C1R, IGFBP3, AHSG, APOA1, ATRN, IGFALS, PRDX1, F9, CRTAC1, TF, LUM, SERPING1 | LR | 0.98 / 0.91 | 0.95 / 0.87 | 0.86 / 0.76 |
|  | RF | 0.96 / 0.86 | 0.94 / 0.88 | 0.90 / 0.89 |
|  | kNN | 0.89 / 0.85 | 0.91 / 0.88 | 0.92 / 0.91 |
|  | SVC | 0.98 / 0.83 | 0.94 / 0.81 | 0.83 / 0.91 |
| **13 proteins:**<br>C1R, IGFBP3, AHSG, APOA1, ATRN, IGFALS, PRDX1, F9, CRTAC1, TF, LUM, SERPING1, SERPINA6 | LR | 0.98 / 0.88 | 0.95 / 0.90 | 0.88 / 0.77 |
|  | RF | 0.95 / 0.88 | 0.94 / 0.86 | 0.94 / 0.93 |
|  | kNN | 0.93 / 0.81 | 0.91 / 0.86 | 0.90 / 0.93 |
|  | SVC | 0.99 / 0.81 | 0.94 / 0.81 | 0.90 / 0.89 |

In order to improve the model’s performance for classifying both mild versus severe patients, as well as controls versus all patients, the protein panel required further improvements. LUM and SERPING1 were appended as they demonstrated the highest *p*-values in the latter comparison. Further evaluation of different combinations of features revealed that addition of TF and SERPINA6 particularly enhanced the classification of different groups achieving the best ROC-AUC (0.94-0.95) and accuracy (0.86-0.93) metrics when using the RF algorithm (Table 3). Therefore, the resulting universal panel consisted of 13 proteins including с1R, IGFBP3, AHSG, APOA1, ATRN, IGFALS, PRDX1, F9, CRTAC1, TF, LUM, SERPING1, and SERPINA6. The RF-13 classifier was found to be the best in different pairwise comparisons and demonstrated a high ROC-AUC >0.9 and accuracy>0.8 for all comparison groups (Fig. 3). This model turned out to be the most versatile among all of the considered, as it well determines both COVID-19-positivity in general, and allows to detect severe and even lethal cases.

**Figure 3.**
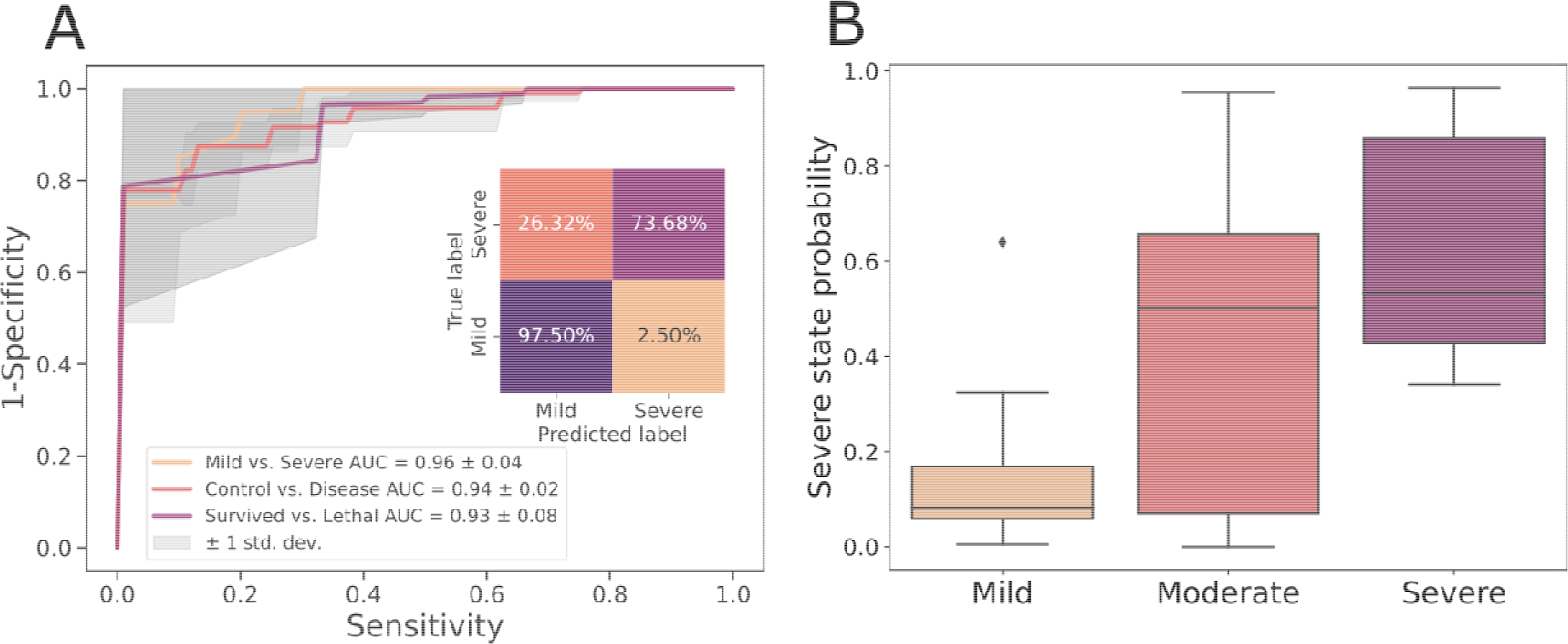
Characteristics of the best performing RF-13 classifier distinguishing COVID-19 patients. (A) ROC-AUC metrics for different pairwise comparisons using 4-fold cross-validation. Insert -Confusion matrix for the classifier prediction of mild or severe COVID-19 course of progression; (B) Probability scores obtained by the developed RF-13 classifier. The scores were obtained by applying the RF-13 classifier to a test set (all moderate patients and 30% of the mild and severe samples) after training (on the rest 70% of the mild and severe samples).

For further validation of the developed RF-13 classifier 70% of the mild/severe patients dataset were used for training, while the remaining 30% and all samples from moderate patients were used as a test set. The resulting probabilities assigned by the classifier to specific samples (Fig. 3B) show a very good agreement with the diagnosed severity of COVID-19.

## Discussion

The obtained results mainly confirm the possibility of using frozen whole blood as an object for MRM analysis of plasma proteins and assessment of the status of patients with COVID-19. Overall, the results are in good agreement with those obtained in MS studies on serum or plasma from COVID-19 patients. In addition, good agreement between measurements in whole blood and plasma samples was demonstrated for most of the targeted proteins, and this result is consistent with a recent study showing high proteomic similarity between plasma and venous or capillary whole blood samples [37]. All of the above is in favor of considering whole blood as an object of proteomic analysis, the relevance of which may increase under conditions of an increased sample influx, since whole blood does not require additional sample preparation and can be collected in small volumes at home by the patient himself.

In general, of the 61 measured reproducible markers, only 5 did not match with any study (FGG, CA1, C1R, S100A9, and F12). Importantly, 9 of the 10 highly reproducible COVID-19 proteomic markers confirmed their significant up-(SERPING1, ORM1, SAA1/SAA2, LBP, CRP, C9) or down-regulation (APOA1, AFM), IGFALS) trends in whole blood samples, in full agreement with the serum and plasma MS studies. Notably, the obtained results showed only minor inconsistencies with these studies (Table 2). Even without taking into account the partial depletion of proteins in some of the other studies (which analyze serum or use depletion of major proteins), it is important to note that in half of the cases, earlier results also have some inconsistencies with each other for a number of proteins, including HP, IGFBP3, SERPINA4, SERPINA7, SERPIND1, C8B and other (Table 2). However, the absolute discrepancy with several other studies still requires further validation on the use of particular proteins as potential markers in whole blood.

The considered variants of marker panels first of all were aimed at distinguishing surviving and lethal patients, and as little as just 9 proteins was sufficient to reach the best combination of ROC-AUC and accuracy metrics of >0.90 with RF and kNN algorithms. Importantly, 5 of these proteins (C1R, IGFBP3, AHSG, APOA1, and IGFALS) turned out to be correlated between whole blood and plasma samples. AHSG demonstrated the best correlation, and together with APOA1 showed significant differences in a number of pairwise comparisons including ‘disease vs. control’, ‘severe vs. mild’, and ‘lethal vs. survived’ (Table 2). Only C1R demonstrated an inconsistent with the other studies dysregulation trend. Among the non-correlating proteins (ATRN, PRDX1, F9, and CRTAC1) PRDX1 was also in the outliers in the regression analysis. Importantly, of the proteins selected for different variants of the panel, only this one was not previously identified as a potential marker of COVID-19 in MS-proteomic studies. It is likely that the poor agreement between plasma and blood measurements of this protein demonstrated in this study may explain why this protein was not significant in plasma and serum studies. However, it is important to note that PRDX2, another protein involved in the same pathways as PRDX1, also turned out to be significantly different in the current study, with regulation consistent with other works and, moreover, with the same upward trend as PRDX1.

Expansion of the panel with proteins that showed the lowest *p*-values in comparisons between controls and patients (SERPING1 and LUM), as well as with additional proteins that distinguish well different groups (TF and SERPINA6), made it possible to obtain a more universal classifier that distinguishes not only survivors from the lethal cases, but also patients from controls and patients by severity. However, to distinguish between particular groups, the composition of the protein panel may be varied to further improve its performance. In particular, discrimination from controls can be improved by monitoring levels of ORM1, F13, APOA4, and APOH. While for distinguishing between severe and mild patients the panel may be enhanced with APOM, FETUB, GC, and SERPINA7.

In general, it seems very important that for most of the analyzed proteins, the results obtained with frozen whole blood samples are consistent with those from plasma and serum. This suggests that some extrapolations are appropriate. However, the results of our study clearly demonstrate that the profiles of important proteins in plasma, serum, and whole blood samples can be somewhat different, and this is important to consider when creating specific diagnostic protein panels.

## Data Availability

All experimental results were uploaded to the PeptideAtlas SRM Experiment Library (PASSEL) and are available via link: http://www.peptideatlas.org/PASS/PASS05842 (accessed on 4 September 2023).

## Acknowledgments

In part of targeted proteomic analysis E.N.N., A.S.K., P.A.S and A.G.B. acknowledge the support by a MegaGrant of the Ministry of Science and Higher Education of the Russian Federation [Agreement with Skolkovo Institute of Science and Technology, #075-10-2022-090 (075-10-2019-083)].

In part of sample preparation and data analysis I.N.K., A.E.B, N.V.Z., M.I.I. are gratefully appreciate funding from the Ministry of Science and Higher Education of the Russian Federation (# 44.1, 44.2 and 44.4).

## References

[1] Geyer, P. E., Holdt, L. M., Teupser, D., & Mann, M. (2017). Revisiting biomarker discovery by plasma proteomics. Molecular systems biology, 13(9), 942.

[2] Whetton, A. D., Preston, G. W., Abubeker, S., & Geifman, N. (2020). Proteomics and informatics for understanding phases and identifying biomarkers in COVID-19 disease. Journal of proteome research, 19(11), 4219–4232.

[3] Bikdeli, B., Madhavan, M. V., Jimenez, D., Chuich, T., Dreyfus, I., Driggin, E., … & Lip, G. Y. (2020). COVID-19 and thrombotic or thromboembolic disease: implications for prevention, antithrombotic therapy, and follow-up: JACC state-of-the-art review. Journal of the American college of cardiology, 75(23), 2950–2973.

[4] Teuwen, L. A., Geldhof, V., Pasut, A., & Carmeliet, P. (2020). COVID-19: the vasculature unleashed. Nature Reviews Immunology, 20(7), 389–391.

[5] Cugno, M., Meroni, P. L., Gualtierotti, R., Griffini, S., Grovetti, E., Torri, A., … & Peyvandi, F. (2020). Complement activation in patients with COVID-19: A novel therapeutic target. Journal of Allergy and Clinical Immunology, 146(1), 215–217.

[6] Vincent, J. L., & Taccone, F. S. (2020). Understanding pathways to death in patients with COVID-19. The Lancet Respiratory Medicine, 8(5), 430–432.

[7] Kermali, M., Khalsa, R. K., Pillai, K., Ismail, Z., & Harky, A. (2020). The role of biomarkers in diagnosis of COVID-19–A systematic review. Life sciences, 254, 117788.

[8] Al-Nesf, M. A., Abdesselem, H. B., Bensmail, I., Ibrahim, S., Saeed, W. A., Mohammed, S. S., … & Al-Ejeh, F. (2022). Prognostic tools and candidate drugs based on plasma proteomics of patients with severe COVID-19 complications. Nature communications, 13(1), 946.

[9] Byeon, S. K., Madugundu, A. K., Garapati, K., Ramarajan, M. G., Saraswat, M., Kumar-M, P., … & Pandey, A. (2022). Development of a multiomics model for identification of predictive biomarkers for COVID-19 severity: a retrospective cohort study. The Lancet Digital Health, 4(9), e632–e645.

[10] Kim, W. Y. (2022). Multi-omic approach to identify risk markers specific to COVID-19. EBioMedicine, 79.

[11] Shen, B., Yi, X., Sun, Y., Bi, X., Du, J., Zhang, C., … & Guo, T. (2020). Proteomic and metabolomic characterization of COVID-19 patient sera. Cell, 182(1), 59–72.

[12] Messner, C. B., Demichev, V., Wendisch, D., Michalick, L., White, M., Freiwald, A., … & Ralser, M. (2020). Ultra-high-throughput clinical proteomics reveals classifiers of COVID-19 infection. Cell systems, 11(1), 11–24.

[13] D’Alessandro, A., Thomas, T., Dzieciatkowska, M., Hill, R. C., Francis, R. O., Hudson, K. E., … & Hansen, K. C. (2020). Serum proteomics in COVID-19 patients: altered coagulation and complement status as a function of IL-6 level. Journal of proteome research, 19(11), 4417–4427.

[14] Di, B., Jia, H., Luo, O. J., Lin, F., Li, K., Zhang, Y., … & Yang, Z. (2020). Identification and validation of predictive factors for progression to severe COVID-19 pneumonia by proteomics. Signal transduction and targeted therapy, 5(1), 217.

[15] Demichev, V., Tober-Lau, P., Nazarenko, T., Thibeault, C., Whitwell, H., Lemke, O., … & Kurth, F. (2020). A time-resolved proteomic and diagnostic map characterizes COVID-19 disease progression and predicts outcome. MedRxiv, 2020-11. doi: 10.1101/2020.11.09.20228015.

[16] Shu, T., Ning, W., Wu, D., Xu, J., Han, Q., Huang, M., … & Zhou, X. (2020). Plasma proteomics identify biomarkers and pathogenesis of COVID-19. Immunity, 53(5), 1108–1122.

[17] Park, J., Kim, H., Kim, S. Y., Kim, Y., Lee, J. S., Dan, K., … & Han, D. (2020). In-depth blood proteome profiling analysis revealed distinct functional characteristics of plasma proteins between severe and non-severe COVID-19 patients. Scientific reports, 10(1), 22418. doi: 10.1038/s41598-020-80120-8.

[18] Geyer, P. E., Arend, F. M., Doll, S., Louiset, M. L., Virreira Winter, S., Müller□Reif, J. B., … & Teupser, D. (2021). High□resolution serum proteome trajectories in COVID□19 reveal patient□specific seroconversion. EMBO Molecular Medicine, 13(8), e14167.

[19] Suvarna, K., Biswas, D., Pai, M. G. J., Acharjee, A., Bankar, R., Palanivel, V., … & Srivastava, S. (2021). Proteomics and machine learning approaches reveal a set of prognostic markers for COVID-19 severity with drug repurposing potential. Frontiers in physiology, 432.

[20] Völlmy, F., Van Den Toorn, H., Chiozzi, R. Z., Zucchetti, O., Papi, A., Volta, C. A., … & Heck, A. J. (2021). A serum proteome signature to predict mortality in severe COVID-19 patients. Life Science Alliance, 4(9).

[21] Wang, Z., Cryar, A., Lemke, O., Tober-Lau, P., Ludwig, D., Helbig, E. T., … & Ralser, M. (2022). A multiplex protein panel assay for severity prediction and outcome prognosis in patients with COVID-19: An observational multi-cohort study. EClinicalMedicine, 49, 101495.

[22] Reverté, L., Yeregui, E., Olona, M., Gutiérrez-Valencia, A., Buzón, M. J., Martí, A., … & COVIDOMICS Study Group. (2022). Fetuin-A, inter-α-trypsin inhibitor, glutamic acid and ChoE (18: 0) are key biomarkers in a panel distinguishing mild from critical coronavirus disease 2019 outcomes. Clinical and Translational Medicine, 12(1), e704.

[23] Mohammed, Y., Goodlett, D. R., Cheng, M. P., Vinh, D. C., Lee, T. C., Mcgeer, A., … & ARBs CORONA I. (2022). Longitudinal plasma proteomics analysis reveals novel candidate biomarkers in acute COVID-19. Journal of proteome research, 21(4), 975–992.

[24] Richard, V. R., Gaither, C., Popp, R., Chaplygina, D., Brzhozovskiy, A., Kononikhin, A., … & Borchers, C. H. (2022). Early prediction of COVID-19 patient survival by targeted plasma multiomics and machine learning. Molecular & Cellular Proteomics, 21(10).

[25] Martin NJ, Bunch J, Cooper HJ (2013) Dried blood spot proteomics: surface extraction of endogenous proteins coupled with automated sample preparation and mass spectrometry analysis. J Am Soc Mass Spectrom 24(8): 1242–1249. 10.1007/s13361-013-0658-1

[26] Chambers AG, Percy AJ, Yang J, Camenzind AG, Borchers CH (2013) Multiplexed quantitation of endogenous proteins in dried blood spots by multiple reaction monitoring-mass spectrometry. Mol Cell Proteomics 12(3):781–791. 10.1074/mcp.M112.022442

[27] Rosting C, Yu J, Cooper HJ (2018) High field asymmetric waveform ion mobility spectrometry in nontargeted bottom-up proteomics of dried blood spots. J Proteome Res 17(6):1997–2004. 10.1021/acs.jproteome.7b00746

[28] Molloy MP, Hill C, O’Rourke MB, Chandra J, Steffen P, McKay MJ, Pascovici D, Herbert BR (2022) Proteomic analysis of whole blood using volumetric absorptive microsampling for precision medicine biomarker studies. J Proteome Res 21(4):1196–1203. 10.1021/acs.jproteome.1c00971

[29] Braudeau, C., Salabert-Le Guen, N., Chevreuil, J., Rimbert, M., Martin, J. C., & Josien, R. (2021). An easy and reliable whole blood freezing method for flow cytometry immuno-phenotyping and functional analyses. Cytometry Part B: Clinical Cytometry, 100(6), 652–665.

[30] Gaither, C., Popp, R., Mohammed, Y., & Borchers, C. H. (2020). Determination of the concentration range for 267 proteins from 21 lots of commercial human plasma using highly multiplexed multiple reaction monitoring mass spectrometry. Analyst, 145(10), 3634–3644.

[31] Kononikhin, A. S., Zakharova, N. V., Semenov, S. D., Bugrova, A. E., Brzhozovskiy, A. G., Indeykina, M. I., … & Nikolaev, E. N. (2022). Prognosis of Alzheimer’s disease using quantitative mass spectrometry of human blood plasma proteins and machine learning. International Journal of Molecular Sciences, 23(14), 7907.

[32] Virtanen, P.,,,,, Gommers, R.,,,,, Oliphant, T.E.,,,,, Haberland, M.,,,,, Reddy, T.,,,,, Cournapeau, D.,,,,, Burovski, E.,,,,, Peterson, P.,,,,, Weckesser, W.,,,,, Bright, J.,,,,, et al. SciPy 1.0: Fundamental algorithms for scientific computing in Python. Nat. Methods 2020, 17, 261–272. 10.1038/s41592-019-0686-2.

[33] Waskom, M.L. Seaborn: Statistical data visualization. J. Open Source Softw. 2021, 6, 3021. 10.21105/joss.03021.

[34] Hunter, J.D. Matplotlib: A 2D graphics environment. Comput. Sci. Eng. 2007, 9, 90–95. 10.1109/MCSE.2007.55.

[35] McKinney, W. Data structures for statistical computing in python. In Proceedings of the 9th Python in Science Conference Austin, TX, USA, 28 June–3 July 2010; Volume 445, pp. 51–56.

[36] Pedregosa, F.,,,,, Varoquaux, G.,,,,, Gramfort, A.,,,,, Michel, V.,,,,, Thirion, B.,,,,, Grisel, O.,,,,, Blondel, M.,,,,, Prettenhofer, P.,,,,, Weiss, R.,,,,, Dubourg, V.,,,,, et al. Scikit-learn: Machine learning in Python. J. Machine Learn. Res. 2011, 12, 2825–2830.

[37] Whelan, S. A., Hendricks, N., Dwight, Z. L., Fu, Q., Moradian, A., Van Eyk, J. E., & Mockus, S. M. (2023). Assessment of a 60-biomarker health surveillance panel (HSP) on whole blood from remote sampling devices by targeted LC/MRM-MS and discovery DIA-MS analysis. medRxiv, 2023-02.

